# Risk Factors for Cardiovascular Disease in Community-Dwelling Older Adults: The Potential Role of Dual Screening for Chronic Kidney Disease and Sarcopenia

**DOI:** 10.64898/2026.03.29.26349633

**Authors:** Takahiro Nishida, Isaki Hanamura, Sumihisa Honda, Ayumi Honda

## Abstract

**Objectives:** Cardiovascular disease (CVD) is a leading cause of mortality and disability in older populations. This study aimed to identify CVD risk factors in community-dwelling older adults and to examine whether frailty-related factors (sarcopenia and nutritional status) interact with chronic kidney disease (CKD).

**Methods:** This cross-sectional study included 307 community-dwelling Japanese adults aged ≥65 years between September 2024 and March 2025. CVD history was assessed based on self-reported physician diagnoses obtained through a structured questionnaire. Lifestyle-related factors included hypertension, diabetes, dyslipidemia, and body mass index (BMI). Frailty-related factors included sarcopenia (Asian Working Group for Sarcopenia 2019 criteria), nutritional status (Mini Nutritional Assessment–Short Form), and physical activity (International Physical Activity Questionnaire–Short Form). CKD was defined using the estimated glomerular filtration rate (eGFR): non-CKD (≥60 mL/min/1.73 m²) and CKD (<60 mL/min/1.73 m²). Multivariable logistic regression identified independent correlates of CVD, and interactions between CKD and frailty-related factors were tested.

**Results:** The prevalence of CVD was 17.9%. Independent correlates included CKD (aOR 5.0), hypertension (aOR 4.0), male sex (aOR 3.1), undernutrition (aOR 2.7), sarcopenia (aOR 2.7), and low physical activity (aOR 2.5). No significant interactions were observed between CKD and sarcopenia (p = 0.70) or nutritional status (p = 0.40).

**Conclusions:** CKD, sarcopenia, undernutrition, and low physical activity were independently associated with CVD, with no interaction between CKD and frailty factors. These findings suggest that integrated management addressing both renal function and frailty-related factors may be important for CVD prevention in older adults.

## Introduction

Cardiovascular disease (CVD), including ischemic and cerebrovascular diseases, is a leading cause of global mortality and disability.^1–3^ Its burden is particularly heavy among older adults, contributing to mortality, functional decline, and long-term care needs.^1–3^ This issue is critical in Japan, which had the world’s highest life expectancy in 2023 (81.1 years for men; 87.1 for women)^4^ and a 29.1% aging rate.^5^ In 2022, cardiovascular and cerebrovascular diseases were the second to fourth leading causes of death for those aged 60–90, following cancer.^6^ Additionally, primary reasons for long-term care included dementia (16.6%), stroke (16.1%), falls/fractures (13.9%), and debility (13.2%),^7^ underscoring the profound impact of these conditions on both mortality and functional independence.^1–3^

Chronic kidney disease (CKD) is highly prevalent among older adults and is a well-established risk factor for CVD.^8–12^ Approximately 30% of Japanese adults aged ≥65 years are estimated to have CKD.^8,13^ Declining kidney function promotes vascular damage through inflammatory and metabolic pathways, thereby increasing cardiovascular risk.^10,12^

In addition to traditional cardiometabolic risk factors such as hypertension, geriatric conditions including frailty and sarcopenia have gained attention as contributors to adverse cardiovascular outcomes.^10,14^ Sarcopenia, characterized by age-related declines in muscle mass, strength, and physical performance, is a key component of frailty and is common among community-dwelling older adults.^15–19^ Malnutrition and physical inactivity further exacerbate this condition, accelerating functional decline and increasing vulnerability to cardiovascular events.^14,17^

The management of older adults with CKD presents a clinical challenge. While protein restriction is recommended to slow CKD progression, adequate protein intake is necessary to maintain muscle mass and prevent sarcopenia.^8,9,20,21^ This creates a nutritional dilemma in which renal protection may conflict with frailty prevention.^9,22^ Recent guidelines from the Japanese Association on Sarcopenia and Frailty (JASF)^23^ reflect this complexity by recommending different protein intake levels based on CKD stage and sarcopenia status (Figure 1). These considerations highlight the need to assess renal function, nutritional status, and muscle health simultaneously.

**Fig. 1.**
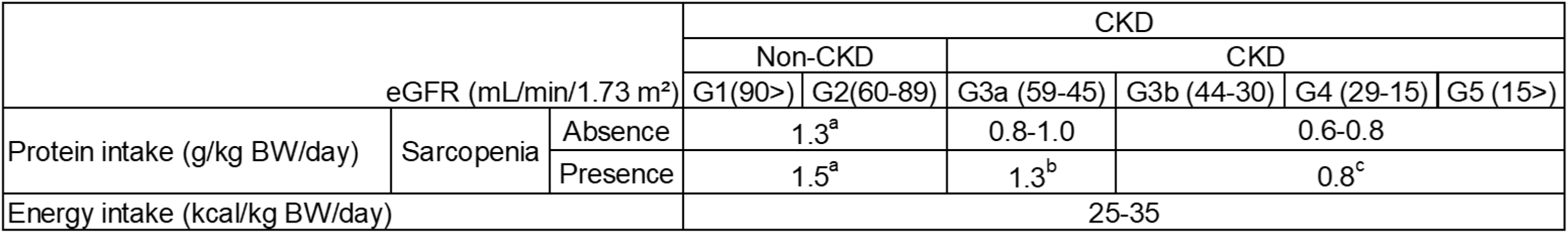
Protein intake considerations and guidelines in the dietary management of CKD with sarcopenia. The Japanese Association on Sarcopenia and Frailty has developed dietary reference values for older adults, recommending appropriate levels of protein and energy intake based on CKD stage and the presence or absence of sarcopenia. ^a^Avoid excessive intake. ^b^In CKD stage G3, patients may require either relaxed or prioritized protein restriction depending on their clinical context. (Relaxed: up to 1.3 g/kg BW/day; Prioritized: upper limit of the recommended intake for the stage). ^c^Protein restriction is generally prioritized but may be relaxed depending on comorbid conditions. (When relaxed: up to 0.8 g/kg BW/day). Note: Relaxed protein restriction should be based on not only GFR and proteinuria, but also factors such as the rate of kidney function decline, absolute risk of ESKD, mortality risk, and degree of sarcopenia. CVD, cardiovascular or cerebrovascular disease; CKD, chronic kidney disease; eGFR, estimated glomerular filtration rate (mL/min/1.73 m^2^); BW, body weight; ESKD, end-stage kidney disease.

Although previous studies have shown that cardiometabolic factors, frailty, and CKD are each associated with CVD risk,^10,12–14^ few have examined their combined effects, particularly the interaction between CKD and frailty-related conditions such as sarcopenia or malnutrition. Therefore, this study aimed to identify risk factors for CVD in community-dwelling older adults, focusing on cardiometabolic conditions, frailty-related factors, and CKD. We also explored the association between protein intake and CVD. We hypothesized that sarcopenia and poor nutritional status would interact with CKD to increase CVD risk.

## Methods

### Study design and participants

This cross-sectional study was conducted from September 2024 to March 2025 in Sasebo City, Nagasaki Prefecture, Japan, as part of a community-based frailty prevention program. Participants were community-dwelling adults aged ≥65 years who attended programs at 48 public community centers. Individuals aged <65 years, those receiving dialysis, those without consent, and those with missing data were excluded. A total of 307 participants were included in the final analysis (Figure 2).

**Fig. 2.**
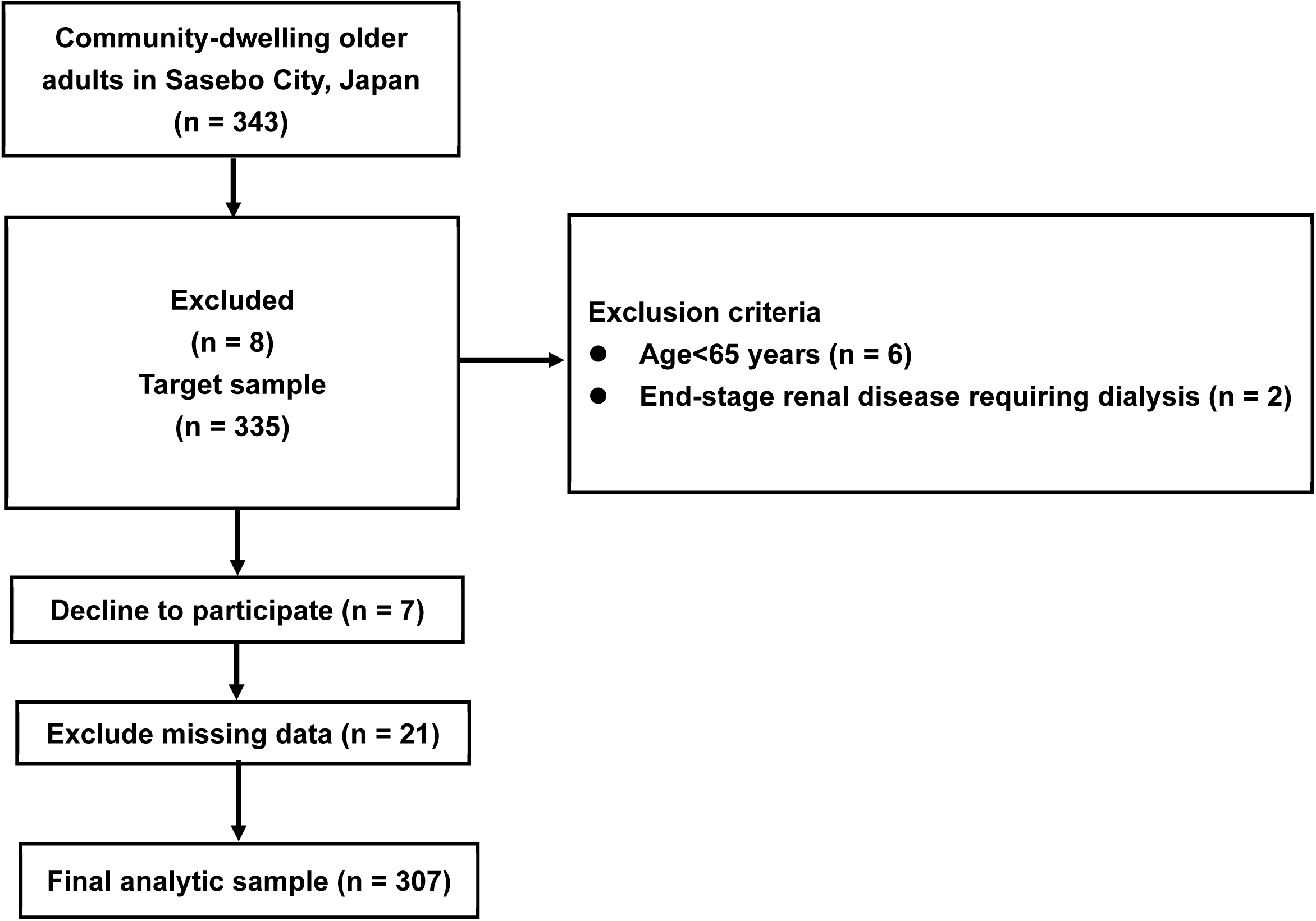
Flowchart of participant selection.

### Data collection and measurements

Participants completed structured questionnaires on demographics (sex, age, living arrangements), lifestyle-related diseases, frailty characteristics, substance use, and dietary preferences. Anthropometric and nutritional assessments were conducted by public health nurses.

### Lifestyle-related diseases

Hypertension, diabetes, and dyslipidemia were defined as current treatment based on physician diagnosis. Obesity was defined as body mass index (BMI) ≥25 kg/m² according to Japanese criteria.^2^

### CKD assessment

Serum creatinine was used to calculate the estimated glomerular filtration rate (eGFR) (mL/min/1.73 m^2^) using the following Japanese Society of Nephrology equations^24^:

- Men: eGFR = 194 × (serum creatinine) ^–1.094 × (age) ^–0.287
- Women: eGFR = 194 × (serum creatinine) ^–1.094 × (age) ^–0.287 × 0.739

According to established guidelines,^25^ eGFR was categorized as G1 (≥ 90), G2 (60–89), G3a (45–59), G3b (30–44), G4 (15–29), or G5 (< 15). Participants were further classified as non-CKD (≥ 60; G1–G2) or CKD (< 60; G3a–G5).

### Frailty-related factors

#### Sarcopenia

Sarcopenia was assessed using the Asian Working Group for Sarcopenia (AWGS) 2019 criteria and the SARC-CalF tool,^19^ which includes five questionnaire items (strength, walking assistance, rising from a chair, stair climbing, and falls history) plus calf circumference (CC; < 34 cm for men, < 33 cm for women). Each questionnaire item was scored 0–2, with CC contributing 10 points if below the cutoff value. The total score ranged from 0 to 20, with scores ≥ 11 indicating possible sarcopenia. Participants with low handgrip strength (< 28 kg for men, < 18 kg for women) and/or impaired lower limb function (five-times chair stand ≥ 12 seconds) were classified as sarcopenic.

#### Nutritional status

Nutritional status was assessed using the Mini Nutritional Assessment–Short Form (MNA-SF),^26^ evaluating six items: appetite, weight loss, mobility, psychological stress/acute illness, neurocognitive status, and BMI (used instead of CC). Total scores (0–14) were categorized as malnourished (0–7), at risk (8–11), and well-nourished (12–14), with lower scores indicating poorer status. For analysis, scores <12 were defined as undernutrition.

#### Physical activity

Physical activity was assessed using the short form of the International Physical Activity Questionnaire (IPAQ-SF).^27^ Weekly time spent performing moderate-to-vigorous-intensity physical activity (MVPA; ≥ 3 metabolic equivalents of task) was calculated, along with total MVPA (minutes/week) and average sedentary time (minutes/day). Participants with < 150 minutes/week of MVPA were classified as having low activity based on the World Health Organization (WHO) guidelines.^28^

### Other variables

Substance use (nonsteroidal anti-inflammatory drugs [NSAIDs], tobacco, alcohol) and dietary salt preference were assessed. NSAID use was classified as current or not. Tobacco use was dichotomized into smokers (current/former) and nonsmokers (never); similarly, alcohol use was divided into drinkers (daily/occasional) and nondrinkers (rarely/never). Salt intake was evaluated using a validated 0–35-point questionnaire,^29^ with scores ≥14 indicating excessive intake.

Protein intake was assessed via the MNA^30^ and meal portion size.^29^ The MNA protein component includes three items: daily dairy (≥1 serving), weekly legumes/eggs (≥2 servings), and daily meat/fish, each scored 1 (“yes”) or 0 (“no”). Portion size was rated on a three-point scale (“less than,” “about the same,” or “more than others”) and multiplied to yield a score (0–9; seven possible levels). Notably, this scoring system provides a semi-quantitative estimate of relative consumption rather than absolute daily protein intake (g/kg/day).

### Primary outcome

The primary outcome was self-reported physician-diagnosed CVD (angina, myocardial infarction, cerebral infarction, or cerebral hemorrhage).

## Statistical analysis

Descriptive statistics were calculated as means (SD) or frequencies (%). Age was categorized as 65–74 or ≥75 years. Group comparisons used t-tests and crude odds ratios (cORs) with 95% confidence intervals (CIs).

Multivariable logistic regression (forced-entry) was used to identify factors associated with CVD. Covariates included sex, age group, hypertension, diabetes, CKD, and low physical activity. Model 1 included sarcopenia, and Model 2 included undernutrition. Adjusted odds ratios (aORs) with 95% CIs were reported.

CVD prevalence across CKD stages was assessed using the Cochran–Armitage trend test, stratified by sarcopenia and nutritional status. Interaction terms between CKD and sarcopenia or nutritional status were tested using logistic regression.

For protein intake analyses, participants were classified into four groups based on CKD and sarcopenia status. Protein intake levels were compared using the Kruskal–Wallis test with Steel–Dwass post hoc analysis.

Protein intake was further dichotomized at the median (score 4) into low and normal intake. Participants were stratified by CKD and protein intake status, and CVD prevalence was compared using chi-square tests. A p-value <0.05 was considered significant.

All analyses were performed using EZR software.^31^

## Results

### Participant characteristics

Of the 307 participants, 55 (17.9%) had a history of CVD: 42 (13.7%) with heart disease (angina or myocardial infarction) and 23 (7.5%) with stroke (infarction or hemorrhage). The mean age was 78.4 years (SD 6.0); 72.0% were aged ≥75 years and 14.3% were male. Hypertension was present in 68.7%, diabetes in 12.4%, dyslipidemia in 46.3%, obesity in 27.4%, and CKD in 52.1%. Among frailty-related factors, 18.6% had sarcopenia, 38.8% were undernourished, and 31.3% had low physical activity.

### Comparison between participants with and without CVD

Compared with participants without CVD, those with CVD had higher prevalences of hypertension (89.1% vs. 64.3%; cOR 4.54, p < 0.01), diabetes (27.3% vs. 9.1%; cOR 3.73, p < 0.01), and CKD (80.0% vs. 46.4%; cOR 4.62, p < 0.01). Frailty-related factors were also more common in the CVD group: sarcopenia (38.2% vs. 14.3%; cOR 3.71, p < 0.01), undernutrition (52.7% vs. 35.7%; cOR 2.01, p = 0.019), and low physical activity (56.4% vs. 25.8%; cOR 3.72, p < 0.01). No significant differences were observed for dyslipidemia or obesity.

### Multivariable logistic regression analysis

Multivariable logistic regression analyses showed that CKD, hypertension, low physical activity, and male sex were consistently associated with CVD in both models (Table 2). In Model 2, independent predictors included CKD (aOR 4.97; 95% CI 2.22–11.10), hypertension (aOR 4.03; 95% CI 1.52–10.70), low physical activity (aOR 2.54; 95% CI 1.27–5.11), and male sex (aOR 3.11; 95% CI 1.35–7.16). Undernutrition in Model 2 (aOR 2.74; 95% CI 1.34–5.58) and sarcopenia in Model 1 (aOR 2.72; 95% CI 1.25–5.95) were also identified as independent predictors. Diabetes showed a borderline association (aOR 2.34; 95% CI 0.99–5.54; p = 0.052).

**Table 1.** Characteristics of the participants by non-CVD and CVD (n = 307).

| Characteristics | Overall<br>(n = 307) | Non-CVD<br>(n = 252) | CVD<br>(n = 55) | cOR | 95% CI | P-value |
| --- | --- | --- | --- | --- | --- | --- |
| **Basic attributes** |  |  |  |  |  |  |
| Sex, male | 44 (14.3) | 29 (11.5) | 15 (27.3) | 2.88 | 1.42–5.86 | 0.003 |
| Age group, old-old | 221 (72.0) | 178 (70.6) | 43 (78.2) | 1.49 | 0.74–2.99 | 0.259 |
| Age, mean (SD) | 78.4 (6.0) | 77.8 (5.8) | 81.2 (6.1) |  |  | <0.001<sup>a</sup> |
| Living arrangement, living alone | 89 (29.0) | 75 (29.8) | 14 (25.5) | 0.81 | 0.42–1.57 | 0.524 |
| **Lifestyle-related disease** |  |  |  |  |  |  |
| Hypertension, presence | 211 (68.7) | 162 (64.3) | 49 (89.1) | 4.54 | 1.87–11.0 | <0.001 |
| Diabetes, presence | 38 (12.4) | 23 (9.1) | 15 (27.3) | 3.73 | 1.80–7.76 | <0.001 |
| Dyslipidemia, presence | 142 (46.3) | 113 (44.8) | 29 (52.7) | 1.37 | 0.77–2.46 | 0.288 |
| Obesity (BMI ≥ 25 kg/m<sup>2</sup>) | 84 (27.4) | 70 (27.8) | 14 (25.5) | 0.89 | 0.46–1.73 | 0.726 |
| **CKD, presence** | 160 (52.1) | 117 (46.4) | 44 (80.0) | 4.62 | 2.28–9.35 | <0.001 |
| **Frailty cycle-related component** |  |  |  |  |  |  |
| Sarcopenia, presence | 57 (18.6) | 36 (14.3) | 21 (38.2) | 3.71 | 1.94–7.09 | <0.001 |
| Nutritional status, undernutrition | 119 (38.8) | 90 (35.7) | 29 (52.7) | 2.01 | 1.11–3.62 | 0.019 |
| MVPA, < 150 (min/week) | 96 (31.3) | 65 (25.8) | 31 (56.4) | 3.72 | 2.03–6.79 | <0.001 |
| MVPA, (total min/week), mean (SD) | 368.6 (449.0) | 398.6 (444.0) | 231.0 (450.2) |  |  | 0.012<sup>a</sup> |
| Sedentary time (min/day), mean (SD) | 382.4 (234.5) | 355.9 (222.2) | 504.0 (252.5) |  |  | <0.001<sup>a</sup> |
| **Substance use and dietary preferences** |  |  |  |  |  |  |
| Salt intake level, excessive | 87 (28.3) | 73 (29.0) | 14 (25.5) | 0.84 | 0.43–1.63 | 0.600 |
| Smoking habit, smoker | 34 (11.1) | 23 (9.1) | 11 (20.0) | 2.49 | 1.13–5.47 | 0.020 |
| Alcohol consumption, drinker | 83 (27.0) | 67 (26.6) | 16 (29.1) | 1.13 | 0.59–2.16 | 0.705 |
| NSAIDs, on medication | 108 (35.2) | 82 (32.5) | 26 (47.3) | 1.86 | 1.03–3.36 | 0.038 |
| **Comorbidities** |  |  |  |  |  |  |
| Anemia | 39 (12.7) | 23 (9.1) | 16 (29.1) | 4.09 | 1.98–8.42 | <0.001 |
| Osteoporosis | 80 (26.1) | 65 (25.8) | 15 (27.3) | 1.08 | 0.56–2.08 | 0.821 |

<sup>a</sup>t-test; cOR, crude odds ratio; CI, confidence interval; CVD, cardiovascular disease; CKD, chronic kidney disease; NSAID, nonsteroidal anti-inflammatory drug; MVPA, moderate-to-vigorous-intensity physical activity; BMI, body mass index.

**Table 2.** Logistic regression analysis of risk factors by CVD (n = 307).

| Variable | Model 1 |  |  | Model 2 |  |  |
| --- | --- | --- | --- | --- | --- | --- |
|  | aOR | 95% CI | P-value | aOR | 95% CI | P-value |
| Sex, male | 3.30 | 1.43–7.61 | 0.005 | 3.11 | 1.35–7.16 | 0.008 |
| Age group, old-old | 0.59 | 0.26–1.37 | 0.219 | 0.63 | 0.28–1.43 | 0.269 |
| **Lifestyle-related disease** |  |  |  |  |  |  |
| Hypertension, presence | 3.10 | 1.19–8.04 | 0.020 | 4.03 | 1.52–10.70 | 0.005 |
| Diabetes, presence | 2.19 | 0.94–5.11 | 0.070 | 2.34 | 0.99–5.54 | 0.052 |
| **CKD, presence** | 4.18 | 1.90–9.15 | <0.001 | 4.97 | 2.22–11.10 | < 0.001 |
| **Frailty cycle-related component** |  |  |  |  |  |  |
| Sarcopenia/ Undernutrition, presence | 2.72 | 1.25–5.95 | 0.012 | 2.74 | 1.34–5.58 | 0.006 |
| MVPA (times/week), < 150 | 2.51 | 1.25–5.04 | 0.010 | 2.54 | 1.27–5.11 | 0.009 |

**Note.** Multivariable logistic regression analysis was performed using the forced-entry method to identify factors associated with cardiovascular disease (CVD).
Model 1 included sarcopenia, whereas Model 2 included nutritional status (undernutrition). Common covariates adjusted in both models were sex, age group (old-old), hypertension, diabetes, chronic kidney disease (CKD), and moderate-to-vigorous-intensity physical activity (MVPA).
aOR, adjusted odds ratio; CI, confidence interval.

### Association between CKD stage and CVD prevalence

CVD prevalence increased with advancing CKD stage, from 0% in G1 to 100% in G5 (p for trend < 0.01) (Figure 3a). This trend remained significant in participants with and without sarcopenia (p < 0.01) (Figure 3b) and in both well-nourished and undernourished groups (p < 0.01) (Figure 3c). No significant interactions were observed between CKD and sarcopenia (p = 0.702) or nutritional status (p = 0.398).

**Fig. 3.**
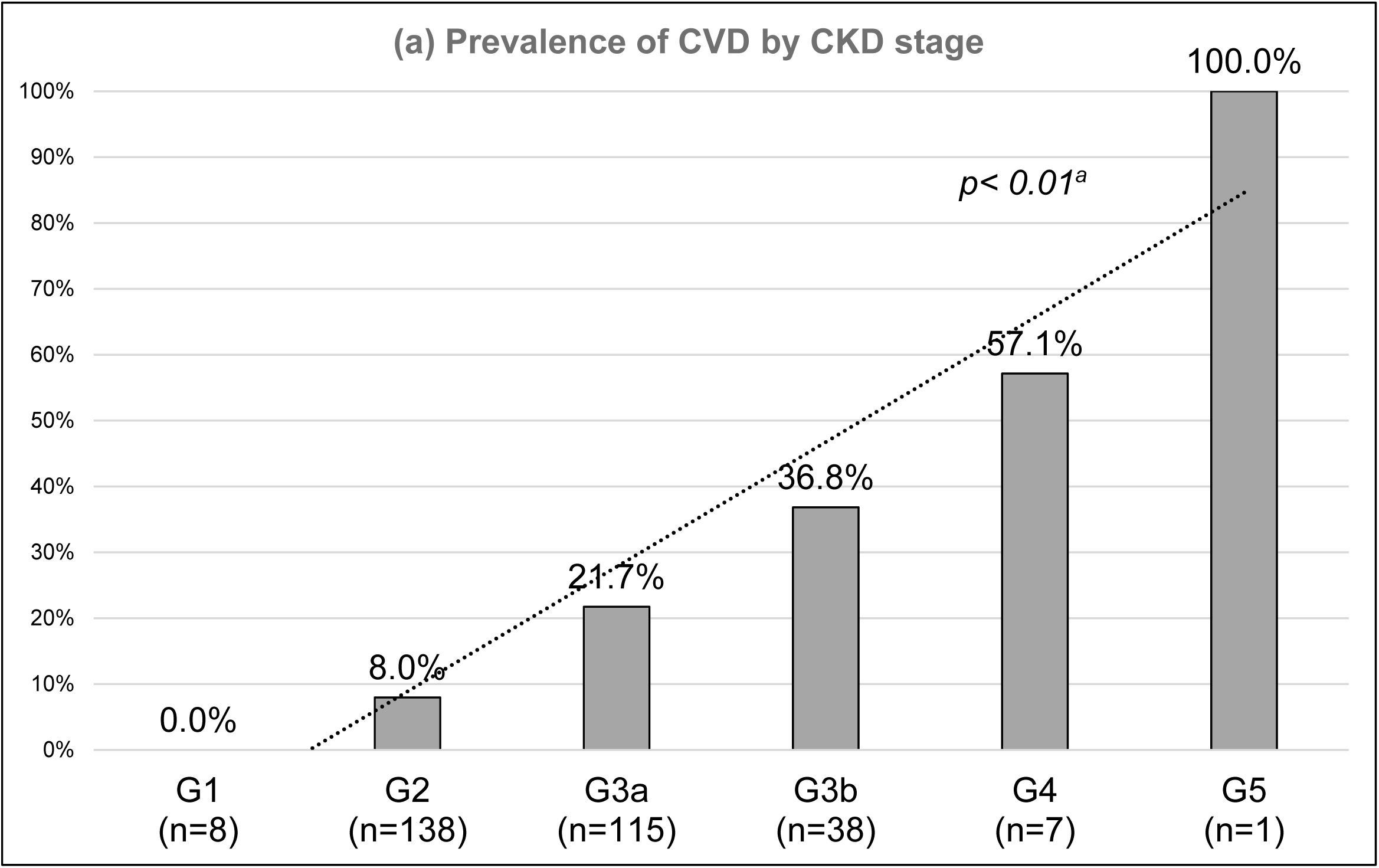
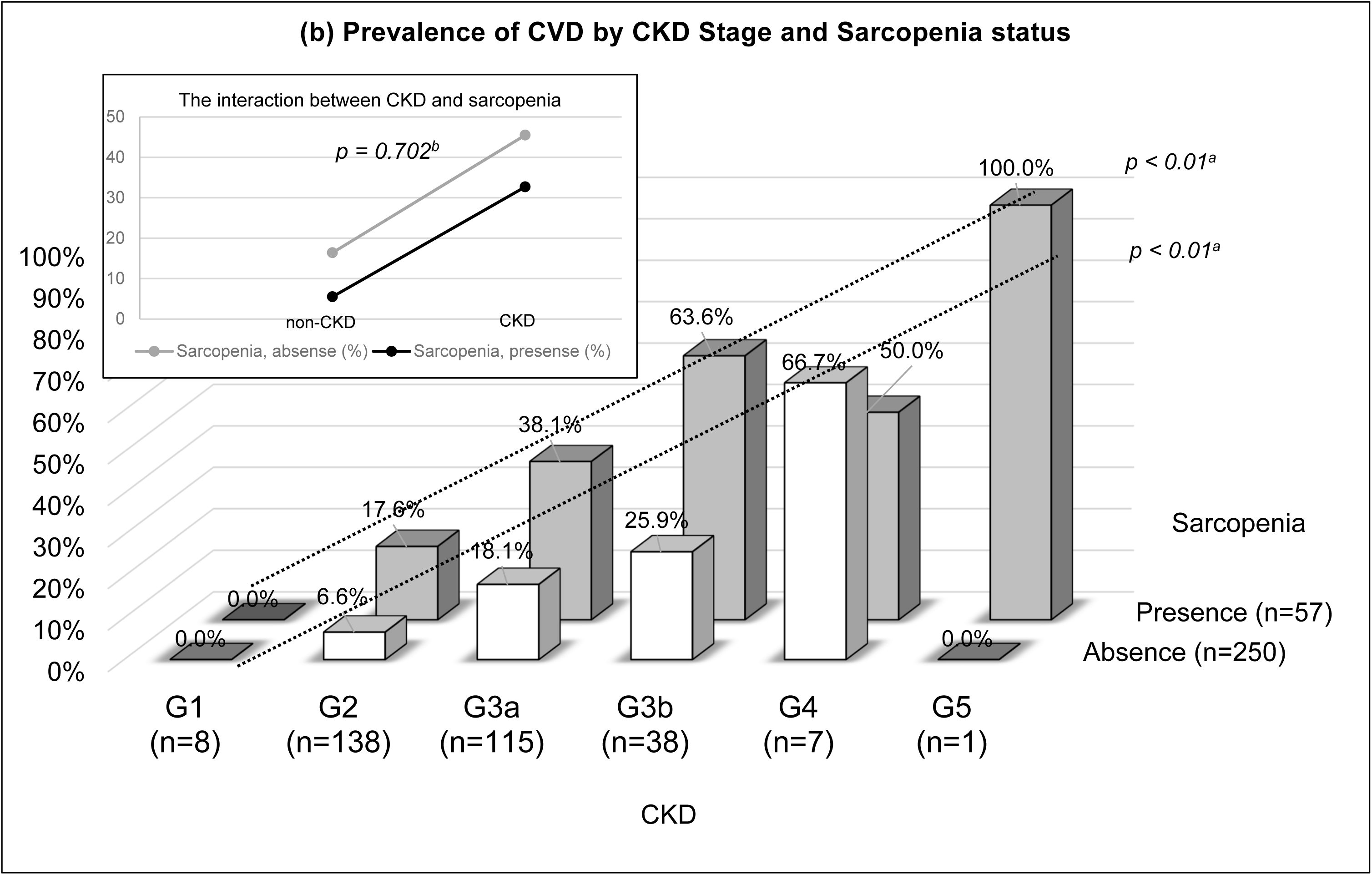
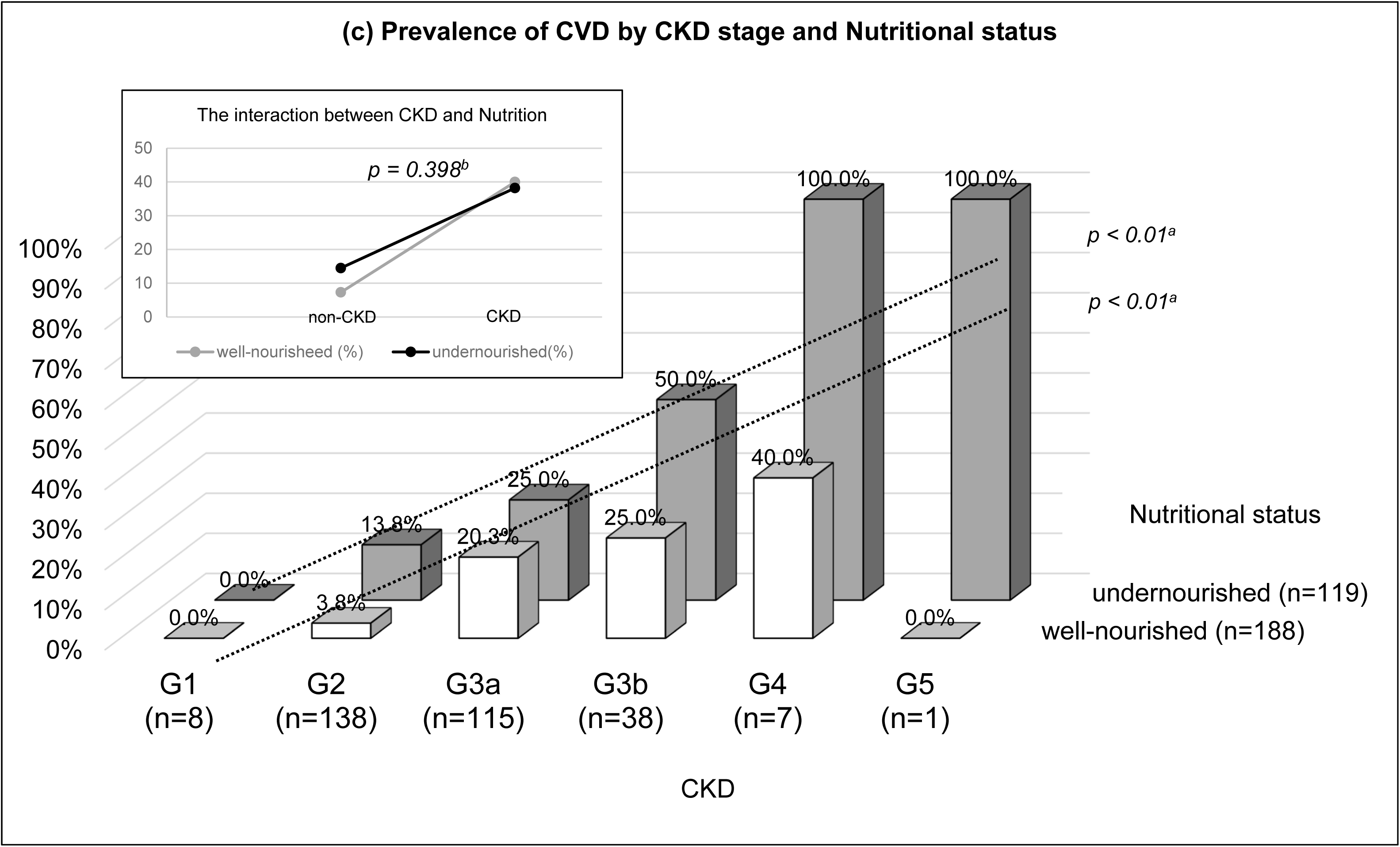
Prevalence of CVD by CKD stage and presence or absence of sarcopenia and malnutrition in community-dwelling older adults (n = 307). (a) Prevalence of CVD by CKD stage. The prevalence of CVD increased progressively with advancing CKD stage (p for trend < 0.01). (b) Prevalence of CVD by CKD stage and sarcopenia status. Similar trends were observed in both the sarcopenic and non-sarcopenic groups (p for trend < 0.01 and p = 0.04, respectively), with no significant CKD × sarcopenia interaction (p = 0.702). (c) Prevalence of CVD by CKD stage and nutritional status. Comparable results were found in the well-nourished and malnourished groups (both p for trend < 0.01), with no significant CKD × nutrition interaction (p = 0.398). CVD, cardiovascular disease; CKD, chronic kidney disease. ^a^Cochran–Armitage test ^b^Interaction effects in logistic regression

### Additional analysis of protein intake

Protein intake differed significantly across groups defined by CKD and sarcopenia status (p < 0.01). Post hoc analysis showed no significant differences between non-CKD participants with and without sarcopenia or within the CKD group (Supplementary Figure 1). When stratified by CKD status and protein intake level, CVD prevalence did not differ by protein intake in the non-CKD group (both 7.5%). In contrast, among participants with CKD, low protein intake was associated with a higher prevalence of CVD compared with normal intake (38.1% vs. 20.4%; p = 0.018) (Supplementary Figure 2).

## Discussion

This study examined factors associated with CVD among community-dwelling older adults, focusing on lifestyle-related diseases, frailty-related conditions, and CKD. Several key findings emerged. First, CKD showed one of the strongest associations with CVD, even after adjusting for established cardiometabolic risk factors. Second, frailty-related conditions, including sarcopenia, undernutrition, and low physical activity, were also associated with CVD. Third, CVD prevalence increased with advancing CKD stage. Finally, no significant interaction was observed between CKD and sarcopenia or nutritional status. However, additional analyses showed that low protein intake among individuals with CKD was associated with a higher prevalence of CVD. These findings suggest that both renal function and frailty-related conditions may play important roles in cardiovascular health among older adults.

### The nutritional paradox: CKD, sarcopenia, and CVD risk

Consistent with previous studies,^12,20^ our results confirm that CVD prevalence increases with CKD stage, identifying CKD as a potent independent risk factor (aOR: 5.0). This association likely stems from systemic vascular vulnerability driven by chronic inflammation, oxidative stress, and mineral dysregulation.^10,32^ Notably, sarcopenia and undernutrition also emerged as independent predictors of CVD, suggesting that frailty-related physiological decline further exacerbates cardiovascular risk in older adults.^14,33^

The co-existence of these conditions creates a “nutritional paradox.”^34^ Previous research has shown that while strict protein restriction (0.6–0.8 g/kg/day) can effectively slow eGFR decline compared with more liberal intake (>0.8 g/kg/day), such restriction in patients aged ≥65 years is associated with higher all-cause mortality over a 4-year follow-up.^22^ This suggests that the renal benefits of protein restriction may be offset by increased vulnerability to sarcopenia, frailty, and protein–energy wasting.^22,34^

Interestingly, we observed no statistically significant interaction between CKD and sarcopenia regarding CVD risk. This lack of synergism may be explained by the aforementioned opposing nutritional requirements, which potentially attenuate each other’s measurable effects. However, our stratified analysis revealed that among participants with CKD, CVD prevalence was significantly higher in those with low protein intake—a pattern not observed in those without CKD. This suggests that excessive protein restriction may inadvertently heighten cardiovascular risk in older CKD patients by aggravating muscle loss. Consequently, recent guidelines increasingly advocate for balancing renal protection with nutritional adequacy, suggesting a relaxation of protein limits when frailty is present.^23,34^ Our findings underscore the necessity of dual screening for CKD and sarcopenia to provide individualized nutritional guidance that prioritizes muscle health alongside renal function to improve cardiovascular outcomes.

### Lifestyle-related risk factors: hypertension and diabetes

Among the factors examined, hypertension was a significant independent risk factor for CVD (aOR: 4.0). Diabetes also showed a strong trend (aOR: 2.3), consistent with extensive evidence highlighting the central role of both conditions in CVD development.^1,20^ In this study, hypertension prevalence among adults aged ≥65 years was approximately 69%, aligning with Japanese national estimates.^35^ Age-related arterial stiffening and cumulative risk-factor exposure^36^ may further increase CVD susceptibility in this population. Moreover, subclinical patterns common in older adults—such as morning surges and masked nocturnal hypertension—are strongly linked to cardiovascular risk.^37,38^ These findings underscore the importance of regular monitoring, including home and 24-hour ambulatory blood pressure measurements.

Regarding diabetes, despite the borderline statistical association, its clinical relevance remains substantial. Chronic hyperglycemia promotes atherosclerosis via oxidative stress, endothelial dysfunction, and advanced glycation end products.^2,39^ Furthermore, diabetes often coexists with sarcopenia, potentially exacerbating insulin resistance and cardiovascular vulnerability.^2,14^ Managing these conditions becomes complex alongside CKD.^40^ For instance, potassium-rich dietary patterns such as the Dietary Approaches to Stop Hypertension diet and the Mediterranean diet may conflict with the need for potassium restriction in advanced CKD to prevent hyperkalemia.^40–42^ These complexities highlight the importance of assessing renal function when implementing lifestyle interventions in older adults.

### Frailty-related risk factors: malnutrition and physical inactivity

Undernutrition (aOR: 2.7) and physical inactivity (aOR: 2.5) were independently associated with CVD. These findings are consistent with previous studies highlighting the role of frailty in cardiovascular health.^33,34^ Undernutrition may increase risk through metabolic and inflammatory pathways and, in severe cases, impair cardiac function.^33^ Furthermore, deficiencies in micronutrients such as iron, folate, and vitamins B_12_ and B_6_ may increase vascular risk through mechanisms including anemia and hyperhomocysteinemia.^32^ Therefore, nutritional strategies should aim to ensure sufficient intake of both macro- and micronutrients. Although the present study did not assess detailed dietary intake, improving dietary variety—such as that evaluated by the Dietary Variety Score^43^—may represent a potential approach to enhancing overall nutritional status in older adults.

Although both CKD stage and nutritional status showed linear associations with CVD, no interaction was observed. This may reflect the complexity of nutritional management in CKD. While we hypothesized a synergistic effect, clinical management may have mitigated this interaction. Alternatively, the effects of CKD and malnutrition may be additive rather than synergistic. These findings support regular assessment of both renal function and nutritional status.

Physical inactivity was another key modifiable risk factor. Participants with <150 min/week of physical activity had a higher prevalence of CVD, consistent with WHO recommendations.^28^ Physical activity improves endothelial function, reduces inflammation, and enhances metabolic control,^13,44–46^ which may explain its protective role. Community-based interventions combining nutrition and physical activity may therefore provide dual benefits.

### Nonsignificant associations: obesity and dyslipidemia

Obesity and dyslipidemia were not significantly associated with CVD. This may reflect the “obesity paradox” observed in older adults.^47^ However, BMI does not adequately capture body composition changes such as sarcopenic obesity, which is associated with adverse outcomes.^2,48^ These findings suggest that muscle mass and body composition may be more informative than BMI alone.

### Sex differences

Male sex was independently associated with CVD (aOR: 3.1). This likely reflects both biological and behavioral factors. Lower estrogen exposure may increase vascular vulnerability,^49^ while lifestyle factors such as smoking and lower daily activity may contribute.^50,51^ Promoting daily physical activity, including routine tasks, may help reduce cardiovascular risk.^44,52^

### Limitations

This study has several limitations. First, the sample was drawn from a single rural region and may not be generalizable to broader populations. Second, the use of volunteers from community care prevention programs introduces potential selection bias. Third, CKD was assessed solely based on the eGFR; the lack of urinalysis reduced the diagnostic precision of the findings. In addition, CVD history was self-reported, which may have led to misclassification. Future studies should incorporate repeated kidney function testing and verify medical histories through chart reviews or physician confirmation. Fourth, the cross-sectional design precludes causal inference; for example, reduced physical activity may represent both a risk factor for and a consequence of CVD. Fifth, although we assessed protein intake, the methodology’s limitations and the relatively small sample size may have reduced the statistical power. Future studies should include larger samples and employ validated quantitative dietary tools, such as the brief-type self-administered diet history questionnaire.^53^ Finally, unmeasured confounders, including psychological stress, sleep quality, socioeconomic status, education, and oral frailty, were not evaluated and thus should be considered in future investigations.

Despite these limitations, this study has important strengths. It comprehensively assessed lifestyle, frailty, and renal factors using validated tools. The finding that CKD was the strongest risk factor highlights the need to integrate renal assessment into frailty-based prevention strategies.

## Conclusion

The findings of the present study indicated that hypertension and diabetes were significant lifestyle-related risk factors for CVD, while undernutrition and low physical activity independently contributed among frailty-related conditions. CKD emerged as a distinct and particularly strong risk factor. While no statistical interaction was found between CKD and sarcopenia or undernutrition, their independent impacts—and the specific risk of low protein intake in CKD—necessitate comprehensive management.

Integrating renal and sarcopenia screening into frailty-prevention programs is essential for tailored nutritional guidance and improved cardiovascular health in older populations.

## Acknowledgments

We are grateful to all the participants for their valuable contributions to this study. The authors thank FORTE Science Communications (https://www.forte-science.co.jp/) for English language editing of the initial manuscript. Additionally, during the revision process, the authors used ChatGPT (OpenAI) to improve the readability and language quality of the newly added sections. After using this tool, the authors reviewed and edited the content as needed and take full responsibility for the final content of the publication.

## Author Contributions

Takahiro Nishida: Conceptualization, Methodology, Investigation, Data curation, Formal analysis, Writing – original draft, Visualization, Supervision, Project administration. Isaki Hanamura: Conceptualization, Writing – review & editing, Supervision. Sumihisa Honda: Conceptualization, Methodology, Formal analysis, Writing – original draft, Writing – review & editing, Visualization, Supervision, Project administration. Ayumi Honda: Conceptualization, Methodology, Formal analysis, Writing – original draft, Writing – review & editing, Visualization, Supervision, Project administration. All authors read and approved the final version of this manuscript.

## Statements and Declarations

### Ethical considerations

This study protocol was reviewed and approved by the Ethics Committee of Nagasaki University Graduate School of Biomedical Sciences (Approval No. 25101401). All procedures were performed in accordance with the ethical standards of the institutional research committee and with the 1964 Declaration of Helsinki and its later amendments.

### Consent to participate

Written informed consent was obtained from all individual participants included in the study.

### Consent for publication

Not applicable.

### Data availability

All data produced in the present study are available upon reasonable request to the authors.

---

**Supplementary Fig. 1.**
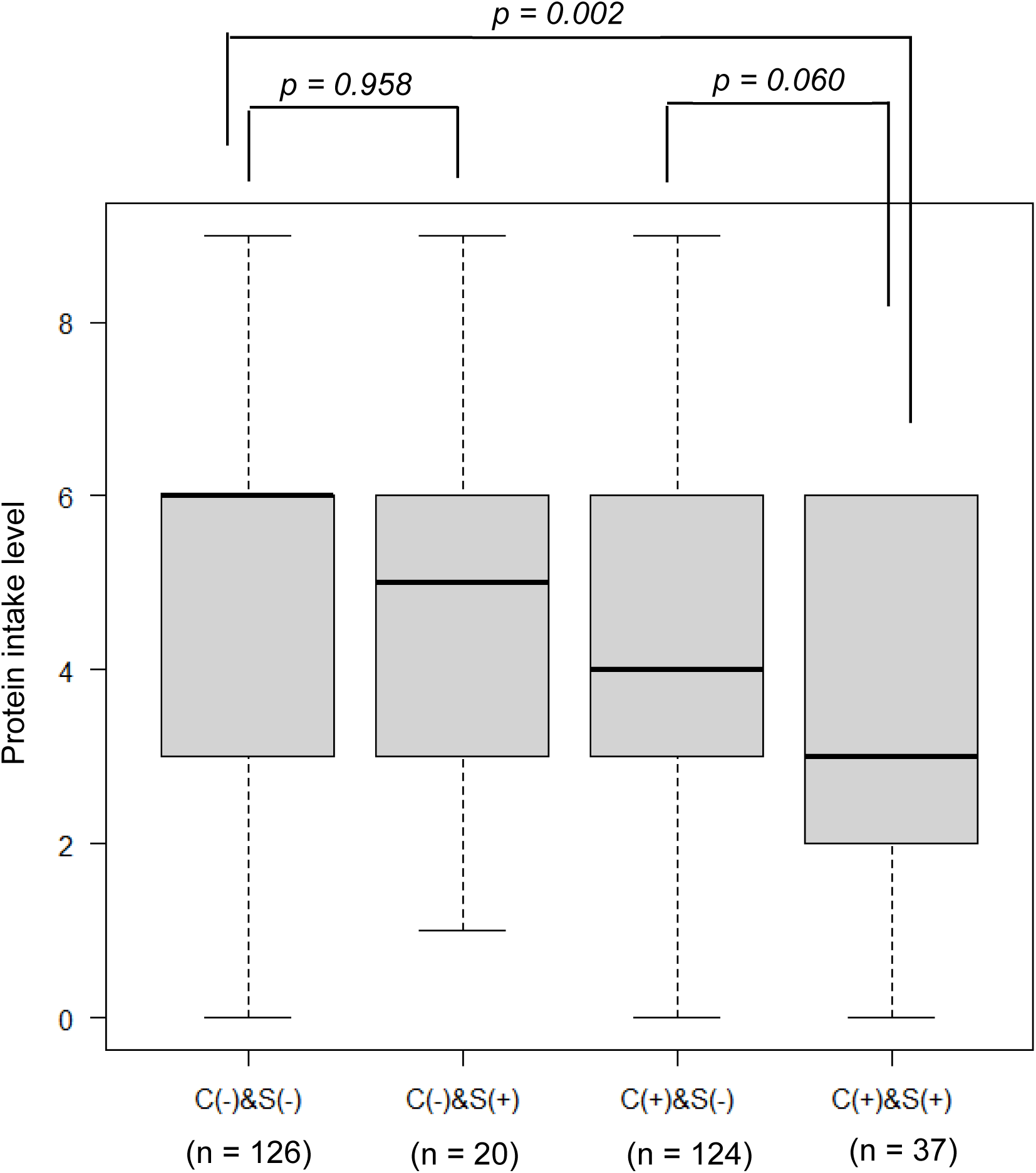
Comparison of protein intake levels among the four groups by CKD and sarcopenia status. Box plots of protein intake scores across four groups defined by CKD and sarcopenia status: C(–)&S(–), C(–)&S(+), C(+)&S(–), and C(+)&S(+). The median [IQR] was 6 [3–6] for C(–)&S(–), 5 [3–6] for C(–)&S(+), 4 [3–6] for C(+)&S(–), and 3 [2–6] for C(+)&S(+). The Kruskal–Wallis test indicated significant differences among groups (p = 0.005), and post hoc analysis showed a significant difference between C(–)&S(–) and C(+)&S(+) (p = 0.002). No other pairwise comparisons were significant. C(+), chronic kidney disease; C(–), non-chronic kidney disease; S(+), presence of sarcopenia; S(–), absence of sarcopenia; IQR, interquartile range.

**Supplementary Fig. 2.**
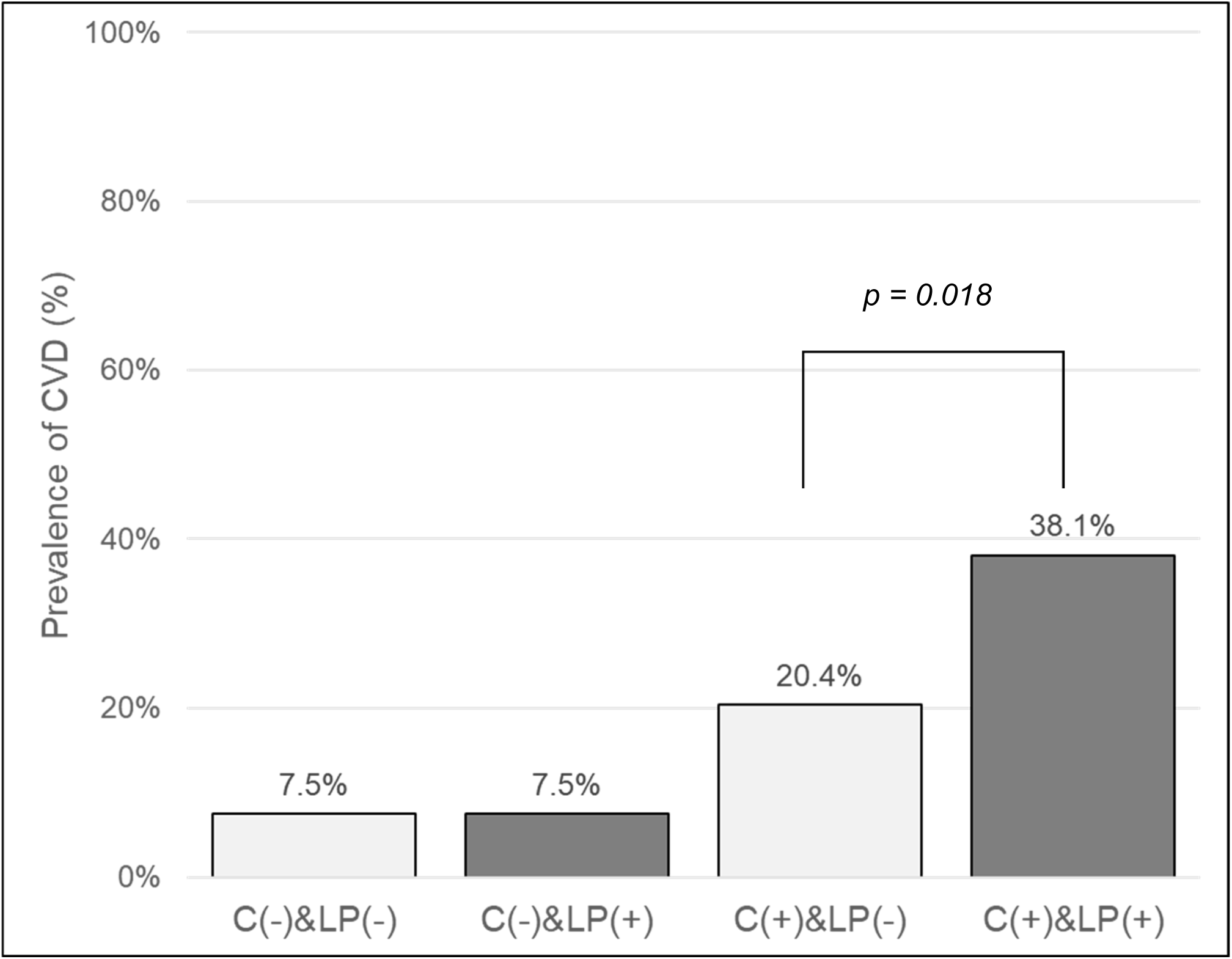
Prevalence of cardiovascular disease according to chronic kidney disease status and protein intake level. Prevalence of cardiovascular disease according to chronic kidney disease status and protein intake level. Participants were stratified into four groups based on the presence of chronic kidney disease (CKD) and protein intake score (dichotomized at the median value of 4). C(−) indicates non-CKD; C(+) indicates CKD. LP(−) indicates normal protein intake (score ≥ 4); LP(+) indicates low protein intake (score < 4). The prevalence of CVD was significantly higher in the low protein intake group compared with the normal protein intake group among participants with CKD (38.1% vs. 20.4%, p = 0.018 by chi-square test). In contrast, no difference was observed in the non-CKD group (7.5% for both). CVD, cardiovascular disease; CKD, chronic kidney disease; LP, low protein intake.

**Supplementary Fig. 3.**
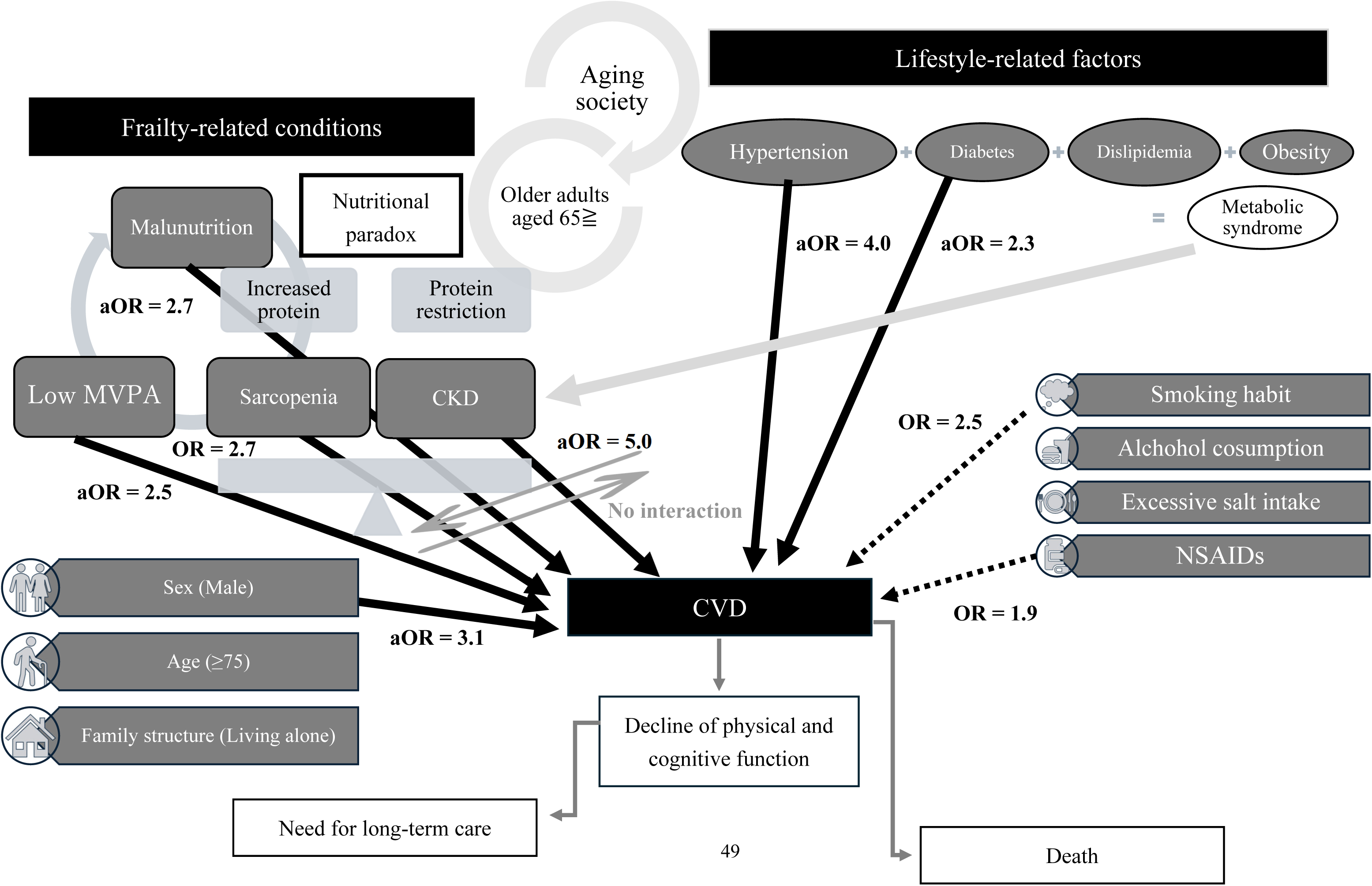
Conceptual study framework and key study findings on risk factors for CVD among community-dwelling older adults. Solid arrows represent the inferred direction of causality, with the strength of the relationship indicated by the adjusted odds ratio. Dashed arrows also represent the inferred direction of causality; however, the strength of this relationship was not significant after adjustment, but was significant in the crude analysis, so the crude odds ratio is reported. The double-bidirectional arrow indicates the absence of an interaction. CKD, chronic kidney disease; CVD, cardiovascular disease; NSAID, nonsteroidal anti-inflammatory drug; MVPA, moderate-to-vigorous-intensity physical activity.

